## Supplementary Material for "Generalizability Challenges of Mortality Risk Prediction Models: A Retrospective Analysis on a Multi-center Database"

**Table of Contents**

**S1 Method.** Difference in Distributions and Statistical Tests

**S2 Method.** Causal Graph Discovery

**S3 Method.** Percentage Change

**S1 Table.** Summary statistics for data after grouping based on hospital ID

**S2 Table.** Summary statistics for data after grouping based on regions

**S3 Table.** List of features

**S4 Table.** Percentage change in metrics from the train hospital to the rest.

**S5 Table.** Percentage change in metrics from the train region to the rest.

**S6 Table.** Description of edge types in causal graph.

**S1 Figure.** Generalization gap in AUC and CS versus dataset shift.

**S2 Figure.** Generalization of performance metrics across regions, number of beds, and teachingstatus.

**S3 Figure.** Causal graph discovered with FCI algorithm.

**S1 Method. Difference in Distributions and Statistical Tests**

To address our objective of explaining lack of external validity, we test for differences in distribution across hospitals and find reasons for the shift. We first require a distance metric to compare datasets and subsequently a procedure to test that the distance is significantly large to label it as a shift. More specifically, we want to compare the distributions from which the hospital datasets are sampled. For any two hospitals $A$ and $B$, say the probability distributions of all measured features are $P_{A}$ and $P_{B}$. Then, we want to estimate a distance metric between $P_{A}$ and $P_{B}$ using samples from the distributions. A widely-used metric is the maximum mean discrepancy (MMD), defined as the distance between mean embeddings of the datasets in a reproducing kernel Hilbert space with a pre-specified kernel (1).

Specifically, it considers a rich class of functions (in a reproducing kernel Hilbert space) and computes the maximum amount by which this function class can separate the two distributions, measured by the maximum difference between the mean of function evaluations at features sampled from each distribution. We chose MMD^2^ since it enables estimating distances between distributions without requiring estimation of their densities, which is a hard task even with moderately small number of features (14 in our case). Additionally, MMD^2^ allows constructing a non-parametric two-sample test to detect whether two datasets differ in distribution (1).An unbiased estimate of MMD^2^ can be computed using samples from the distributions, x_1_*,...,*x*_m_* ∼$P_{A}$ and z_1_*,...,*z*_n_* ∼$P_{B}$ as,

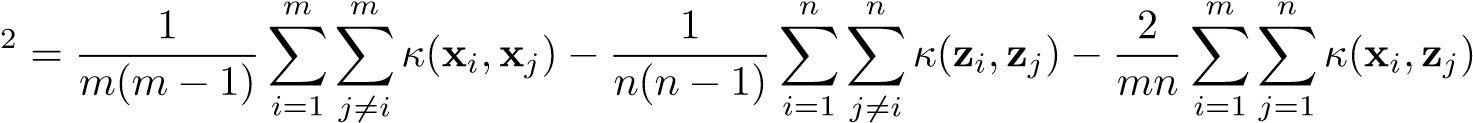

where *κ* denotes the kernel which we take as the Gaussian kernel with width equal to the median Euclidean distance between examples after combining the two datasets. Note that the quantity computed above is an unbiased estimate of MMD^2^, hence it can be negative as well when the true value is close to zero.

Next we want a procedure to test whether $P_{A}$ = $P_{B}$. A permutation test can be performed by noting that under the null hypothesis ($P_{A}$ = $P_{B}$) the dataset examples are exchangeable. This will yield a p-value which can be compared with a desired statistical significance level, fixed as 0.05 here, to decide on the hypothesis.

**S2 Method. Joint Causal Inference and Causal Graph Discovery**

A causal graph encodes relationships between the variables (features and outcomes) via directed edges. An edge X1 → X2 between any two variables X1 and X2 represents that X1 causes X2. For example, temperature and heart rate may be connected in the graph since thermoregulation is intimately tied to metabolic demands of the body, which govern heart rate via a hypothalamic control mechanism. The causal edges are normally informed by domain expertise or, under assumptions about the data generating process, can be discovered from a given dataset. Joint Causal Inference (JCI) aims to represent causal relations for multiple datasets (for example, one for each hospital or region). Differences in relationships between the same variables across datasets are highlighted by adding another set of exogenous variables, which are chosen by the modeler. In our case these are indicators for the hospital and for the geographic region to which each data point belongs. From the discovered graph, we find the direct effects of the indicators, i.e. variables with an edge from indicator (for example, the variables X1, X2 for edges I → X1, I → X2). We interpret such variables as the primary causes for the dataset shift because it is fair to assume that there are no fundamental biological idiosyncrasies between facilities. Due to the causal interpretation of the discovered graph, if these variables did not change across the datasets, the distributions across hospitals or regions will be the same. That is, if X1 and X2 are the only variables directly caused by I, controlling them inhibits any change in I to propagate to other variables except X1 and X2.

We consider the features from the SAPS II risk scoring model for discovering the causal graph. A single causal graph is obtained by pooling together all the data from 179 hospitals. The causal graph is the output of the Fast Causal Inference (FCI) algorithm (2). FCI is a widely used algorithm for causal structure discovery. It uses conditional independence tests to determine graph adjacencies and discover the graph skeleton and then uses a set of orientation rules to determine where to place the edge marks. In accordance with the Joint Causal Inference (JCI) framework (3), the context/exogenous variables used as indicators for hospitals or regions follow the assumptions that no system variable causes any context variable and no context variable is confounded with a system variable. Thus, any adjacent pair of a context variable *k* ∈ *K* and a system variable *i* ∈ *I* is only connected by a directed edge *k* → *i* (since we are assuming no selection bias). Hence, after the adjacency phase of FCI, all edges between a context and a system variable are oriented as *k* → *i*, pointing from the context variable *k* ∈ *K* to the system variable *i* ∈ *I*.

We use the pycausal library^[1]^ in Python built on top of the Tetrad package^[2]^ for learning the causal graph using FCI. The obtained causal graph with all 179 hospitals is shown in S3 Figure. We present details about the edge marks in the causal graph in Table S6.

**S3 Method. Percentage Change**

When transferring from train to test datasets, percentage change in a metric is computed by the following formulae, as done in Wessler et al (4),

$$\frac{(test\_value-baseline)-(train\_value-baseline))}{(train\_value-baseline)} \times100$$

Here, $baseline$ is 0.5 for AUC, and 0 for the other metrics. Variables $test\_value, train\_value$are the mean values of the metrics over the 100 random samples of test and train sets. Percentage change is reported in Table S4 and Table S5.

| Statistics | | | | | | | | | | | Hospital ID | **S1 Table. Summary statistics for data after grouping based on hospital ID.** Table generated with tableone^5^ |
| --- | --- | --- | --- | --- | --- | --- | --- | --- | --- | --- | --- | --- |
|  | race majority, No. (%) |  | is female, No. (%) |  | elective surgery, No. (%) | hosp length of stay, median  [25^th^, 75^th^ percentile] | age, median  [25^th^, 75^th^ percentile] |  | death, No. (%) | n (stays) |  |  |
| False | True | False | True | False | True |  |  | False | True |  |  |  |
| 685 (22.8) | 2320 (77.2) | 1648 (54.8) | 1357 (45.2) | 1945 (64.7) | 1060 (35.3) | 5.9  [3.3,9.6] | 64.0 [53.0,75.0] | 2891 (96.2) | 114 (3.8) | 3005 | ID 73 |  |
| 239 (8.4) | 2594 (91.6) | 1485 (52.4) | 1348 (47.6) | 2485 (87.7) | 348 (12.3) | 5.3  [3.1,8.9] | 63.0 [51.0,74.0] | 2667 (94.1) | 166 (5.9) | 2833 | ID 264 |  |
| 74 (3.2) | 2246 (96.8) | 1314 (56.6) | 1006 (43.4) | 1880 (81.0) | 440 (19.0) | 5.5  [3.4,9.0] | 63.0 [50.0,74.0] | 2192 (94.5) | 128 (5.5) | 2320 | ID 338 |  |
| 1198 (52.5) | 1082 (47.5) | 1310 (57.5) | 970 (42.5) | 1705 (74.8) | 575 (25.2) | 6.0  [3.3,10.7] | 56.0 [42.0,67.0] | 2045 (89.7) | 235 (10.3) | 2280 | ID 443 |  |
| 613 (29.2) | 1489 (70.8) | 1168 (55.6) | 934 (44.4) | 1689 (80.4) | 413 (19.6) | 6.0  [3.4,10.1] | 62.0 [51.0,73.0] | 1915 (91.1) | 187 (8.9) | 2102 | ID 458 |  |
| 98 (5.1) | 1839 (94.9) | 1142 (59.0) | 795 (41.0) | 1604 (82.8) | 333 (17.2) | 5.8  [3.8,9.5] | 64.0 [53.0,75.0] | 1730 (89.3) | 207 (10.7) | 1937 | ID 420 |  |
| 140 (7.4) | 1758 (92.6) | 1091 (57.5) | 807 (42.5) | 1373 (72.3) | 525 (27.7) | 5.3  [3.0,9.5] | 64.0 [53.0,74.0] | 1712 (90.2) | 186 (9.8) | 1898 | ID 252 |  |
| 229 (12.3) | 1628 (87.7) | 1017 (54.8) | 840 (45.2) | 1461 (78.7) | 396 (21.3) | 4.9  [3.1,8.1] | 62.0 [49.0,74.0] | 1763 (94.9) | 94 (5.1) | 1857 | ID 300 |  |
| 429 (23.5) | 1394 (76.5) | 968 (53.1) | 855 (46.9) | 1480 (81.2) | 343 (18.8) | 5.4  [3.4,9.5] | 64.0 [53.0,74.0] | 1679 (92.1) | 144 (7.9) | 1823 | ID 122 |  |
| 485 (29.7) | 1146 (70.3) | 947 (58.1) | 684 (41.9) | 1196 (73.3) | 435 (26.7) | 5.1  [3.2,8.2] | 63.0 [52.0,73.0] | 1491 (91.4) | 140 (8.6) | 1631 | ID 243 |  |

| **S2 Table. Summary statistics for data after grouping based on regions.** Table generated with tableone (5) | | | | | |
| --- | --- | --- | --- | --- | --- |
| **Statistic** |  | **Grouped by Region** | | | |
|  |  | **Midwest** | **Northeast** | **South** | **West** |
| n (stays) |  | 26294 | 5300 | 25181 | 10764 |
| death, No. (%) |  |  |  |  |  |
|  | False | 24799 (94.3) | 4711 (88.9) | 23210 (92.2) | 9766 (90.7) |
|  | True | 1495 (5.7) | 589 (11.1) | 1971 (7.8) | 998 (9.3) |
| age, median  [25^th^, 75^th^ percentile] |  | 64.0 [52.0,75.0] | 64.0 [52.0,75.0] | 64.0 [52.0,75.0] | 64.0 [52.0,75.0] |
| hosp length of stay, median  [25^th^, 75^th^ percentile] |  | 5.2  [3.1,8.8] | 5.7  [3.3,9.2] | 5.4  [3.2,9.0] | 5.1  [3.0,8.6] |
| elective surgery, No. (%) |  |  |  |  |  |
|  | False | 20534 (78.1) | 4503 (85.0) | 20477 (81.3) | 8126 (75.5) |
|  | True | 5760 (21.9) | 797 (15.0) | 4704 (18.7) | 2638 (24.5) |
| is female, No. (%) |  |  |  |  |  |
|  | False | 14469 (55.0) | 3012 (56.8) | 13622 (54.1) | 6003 (55.8) |
|  | True | 11825 (45.0) | 2288 (43.2) | 11559 (45.9) | 4761 (44.2) |
| race majority, No. (%) |  |  |  |  |  |
|  | False | 2972 (11.3) | 291 (5.5) | 6768 (26.9) | 1443 (13.4) |
|  | True | 23322 (88.7) | 5009 (94.5) | 18413 (73.1) | 9321 (86.6) |

| **S3 Table.** **List of features.** Refer to Appendix B in Johnson et al (6) for description of features | | |
| --- | --- | --- |
| **Group** | **Extracted Values** | **Features** |
| Demographics | - | is female, age,  race african american, race hispanic, race asian, race majority (i.e. NOT (race african american, race hispanic, race asian)) |
| SAPS II-derived features | Worst values based on SAPS II scoring sheet in Table 3 of Le Gall et al (7) | [Vitals] heart rate, sysbp (systolic blood pressure), temp (temperature), bg pao2fio2ratio (Oxygen saturation), urine output, GCS (Glasgow coma scale),  [Labs] BUN (blood urea nitrogen), sodium, potassium, bicarbonate, bilirubin, WBC (white blood cell count),  [Demographics, Others] age, elective surgery |
| Outcome | - | death |

| **S4 Table. Percentage change in metrics from the train hospital to the rest.** For each train hospitals and a metric, median, 25th, and 75th percentile are reported for test metrics at the 9 remaining hospitals. Absolute values of DisparityFNR and DisparityCS are used for this analysis. | | | | |
| --- | --- | --- | --- | --- |
| **Metric** | **Hospital ID** | **Median (%)** | **First Quartile (%)** | **Third Quartile (%)** |
| CS | 73 | -45.41 | -57.02 | -39.15 |
| CS | 264 | -19.00 | -22.65 | -8.49 |
| CS | 338 | -15.92 | -18.86 | -7.02 |
| CS | 443 | 17.65 | -0.43 | 24.87 |
| CS | 458 | -20.38 | -25.98 | -6.21 |
| CS | 420 | -26.76 | -34.74 | -17.45 |
| CS | 252 | 2.53 | -5.19 | 10.01 |
| CS | 300 | -22.77 | -25.38 | -17.61 |
| CS | 122 | -16.53 | -22.87 | -4.91 |
| CS | 243 | -25.84 | -33.61 | -17.54 |
| DisparityFNR | 73 | 113.97 | 49.61 | 247.95 |
| DisparityFNR | 264 | -67.87 | -73.21 | -12.36 |
| DisparityFNR | 338 | -72.58 | -78.33 | -62.84 |
| DisparityFNR | 443 | 27.61 | -17.51 | 81.65 |
| DisparityFNR | 458 | -15.93 | -43.82 | 50.08 |
| DisparityFNR | 420 | -58.67 | -61.40 | -29.03 |
| DisparityFNR | 252 | -35.03 | -43.09 | 80.41 |
| DisparityFNR | 300 | 42.87 | -8.31 | 74.07 |
| DisparityFNR | 122 | 187.17 | 72.43 | 278.92 |
| DisparityFNR | 243 | 22.52 | -0.16 | 49.42 |
| DisparityCS | 73 | 59.24 | 9.28 | 135.90 |
| DisparityCS | 264 | -32.67 | -71.83 | 1.16 |
| DisparityCS | 338 | – | – | – |
| DisparityCS | 443 | 102.06 | 48.19 | 158.56 |
| DisparityCS | 458 | 68.03 | 16.41 | 179.21 |
| DisparityCS | 420 | -99.26 | -99.53 | -98.90 |
| DisparityCS | 252 | -52.41 | -70.54 | 75.62 |
| DisparityCS | 300 | 16.46 | -8.93 | 144.37 |
| DisparityCS | 122 | 8.38 | -18.37 | 120.78 |
| DisparityCS | 243 | 17.21 | -27.50 | 91.88 |
| AUC | 73 | -31.45 | -33.73 | -27.21 |
| AUC | 264 | -11.62 | -17.34 | -3.93 |
| AUC | 338 | -4.16 | -8.92 | 6.70 |
| AUC | 443 | -2.54 | -14.96 | 5.79 |
| AUC | 458 | -11.07 | -17.15 | 4.49 |
| AUC | 420 | -20.07 | -25.96 | -11.31 |
| AUC | 252 | 7.95 | 1.28 | 13.50 |
| AUC | 300 | -16.58 | -22.18 | -6.80 |
| AUC | 122 | -11.52 | -16.51 | -8.67 |
| AUC | 243 | -16.73 | -21.49 | -9.61 |

| **S5 Table. Percentage change in metrics from the train region to the rest.** For each train region and a metric, median, 25th, and 75th percentile are reported for test metrics at the 3 remaining regions. Absolute values of DisparityFNR and DisparityCS are used for this analysis. | | | | |
| --- | --- | --- | --- | --- |
| **Metric** | **Region** | **Median (%)** | **First Quartile (%)** | **Third Quartile (%)** |
| CS | South | -5.48 | -8.73 | -2.43 |
| CS | Midwest | -10.48 | -13.33 | -10.38 |
| CS | West | -1.97 | -5.66 | 1.26 |
| CS | Northeast | 1.99 | 0.76 | 5.24 |
| DisparityFNR | South | 37.15 | 14.28 | 43.38 |
| DisparityFNR | Midwest | -17.54 | -28.36 | 4.41 |
| DisparityFNR | West | -32.88 | -38.93 | -11.24 |
| DisparityFNR | Northeast | 64.60 | 30.86 | 108.18 |
| DisparityCS | South | 14.99 | 6.12 | 32.23 |
| DisparityCS | Midwest | -28.76 | -34.44 | -11.31 |
| DisparityCS | West | 67.38 | 31.84 | 88.51 |
| DisparityCS | Northeast | -52.46 | -57.39 | -46.90 |
| AUC | South | -1.52 | -3.76 | -0.02 |
| AUC | Midwest | -8.47 | -9.23 | -7.37 |
| AUC | West | -4.47 | -5.73 | -1.74 |
| AUC | Northeast | -0.81 | -1.72 | 1.00 |

| **S6 Table.** **Description of edge types and the relations exhibited by nodes in the discovered causal graph.** Description is taken from <https://cmu-phil.github.io/tetrad/manual/> | |
| --- | --- |
| Edge Type | Relationships that are present |
| A −− *>* B | A is a cause of B. It may be a direct or indirect cause that may include other measured variables.  Also, there may be an unmeasured confounder of A and B. |
| A *<* − *>* B | There is an unmeasured variable (call it L) that is a cause of A and B. There may be measured variables along the causal pathway from L to A or from L to B. |
| A ◦− *>* B | Either A is a cause of B, or there is an unmeasured variable that is a cause of A and B, or both. |
| A ◦−◦ B | Exactly one of the following holds: (a) A is a cause of B, or  (b) B is a cause of A, or (c) there is an unmeasured variable that is a cause of A and B, or (d) both a and c, or (e) both b and c. |

**S1 Figure. Generalization gap in AUC and CS versus dataset shift measured in MMD^2^ for top 10 hospitals.** Although the test performance worsens in relation to train performance (since magnitude of gap decreases) with increasing dataset shift, the correlation coefficient is not high. Each point is the average gap over 100 random subsamples for each of the 10x10 train-test hospital pairs. Abbreviations: AUC, area under ROC curve; CS, calibration slope.

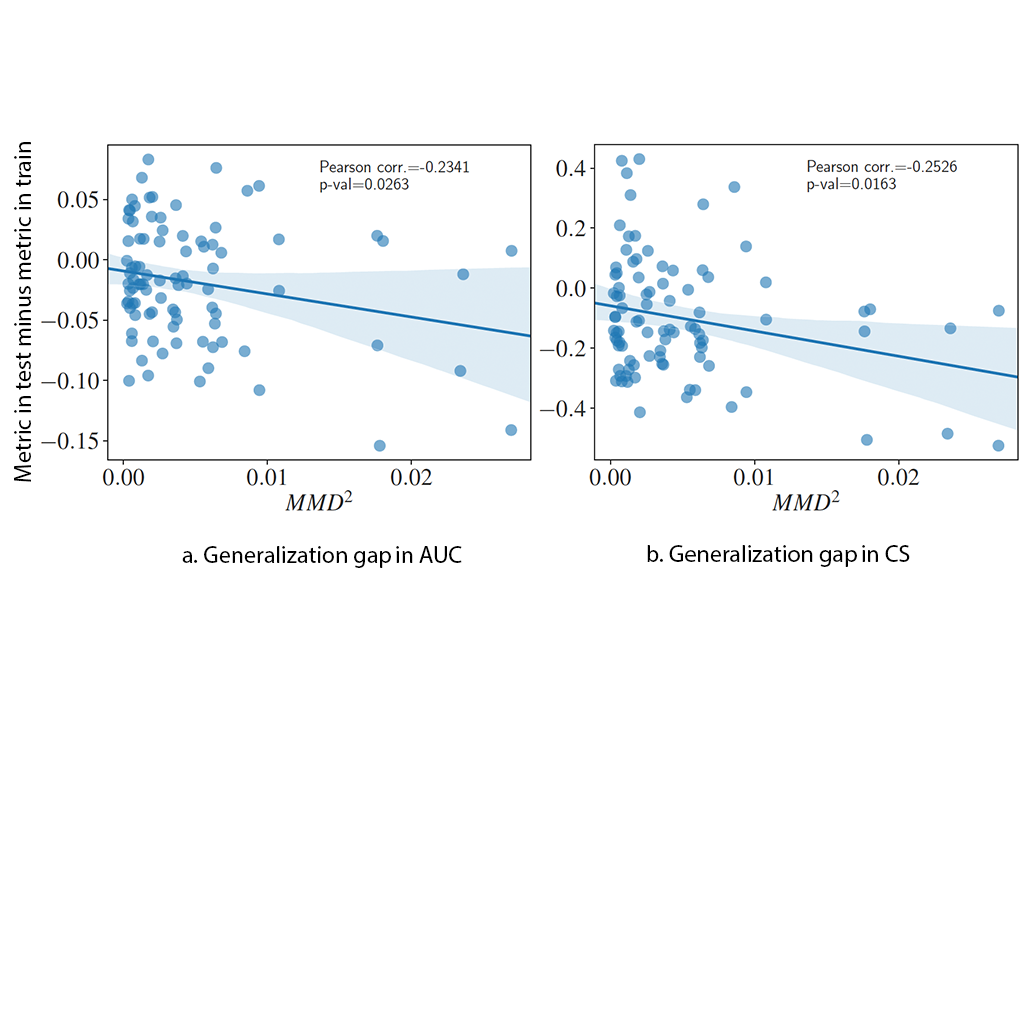

**S2 Figure.** **Generalization of performance metrics across regions, number of beds, and teaching status.** Results of transferring models after pooling hospitals based on region (South, Midwest, West, Northeast), number of beds (less than 100, 100-249, 250-499, more than 500) and teaching status (yes, no). Models are trained and tested on 4000 samples from each category of hospitals. Categories with less than 4000 samples after pooling are excluded from this analysis. DisparityFNR and DisparityCS show large variability when transferring models. Abbreviations: AUC, area under ROC curve; CS, calibration slope; FNR, false negative rate; nbed, number of beds; teach, hospitals with teaching status; noteach, hospitals without teaching status.
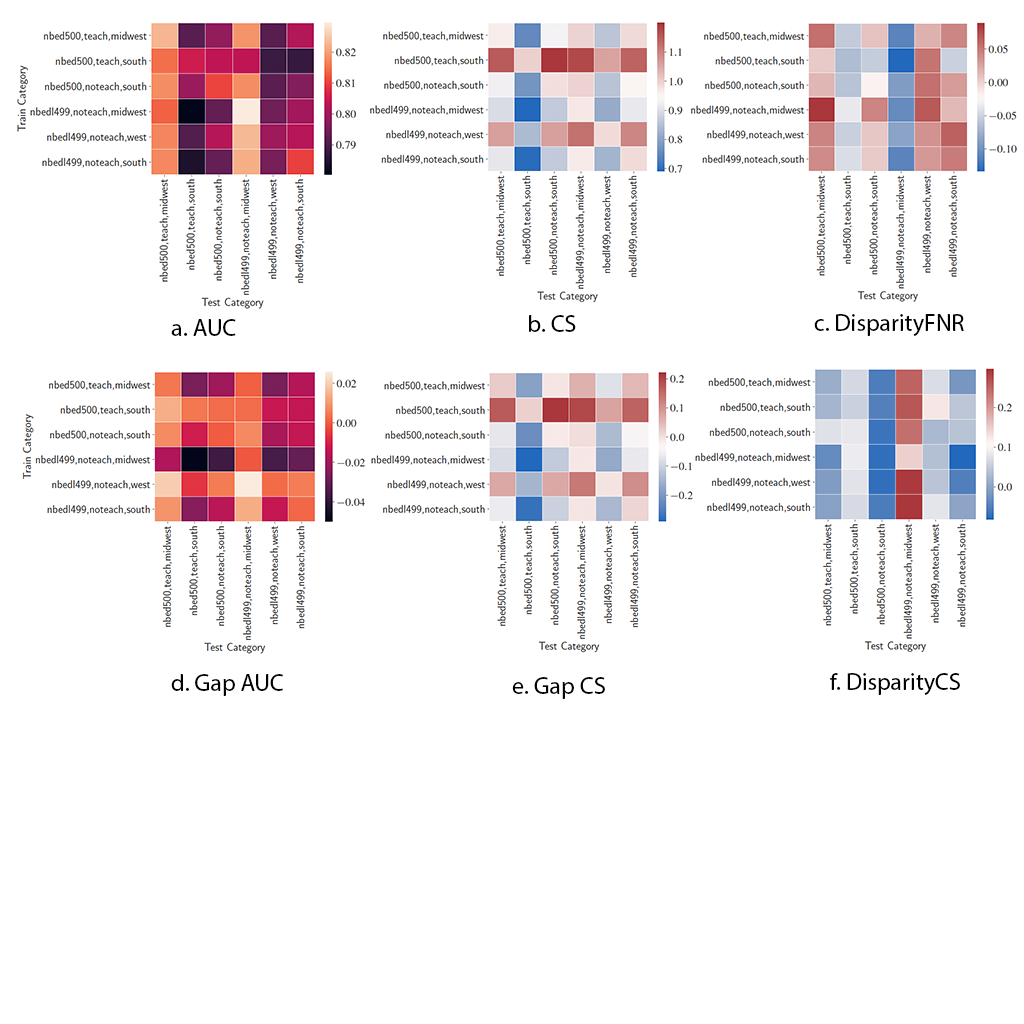

**S3 Figure. Causal graph discovered by the FCI algorithm** (2) **for all hospitals.** Nodes include SAPS II features, race, number of beds, region, and teaching status.

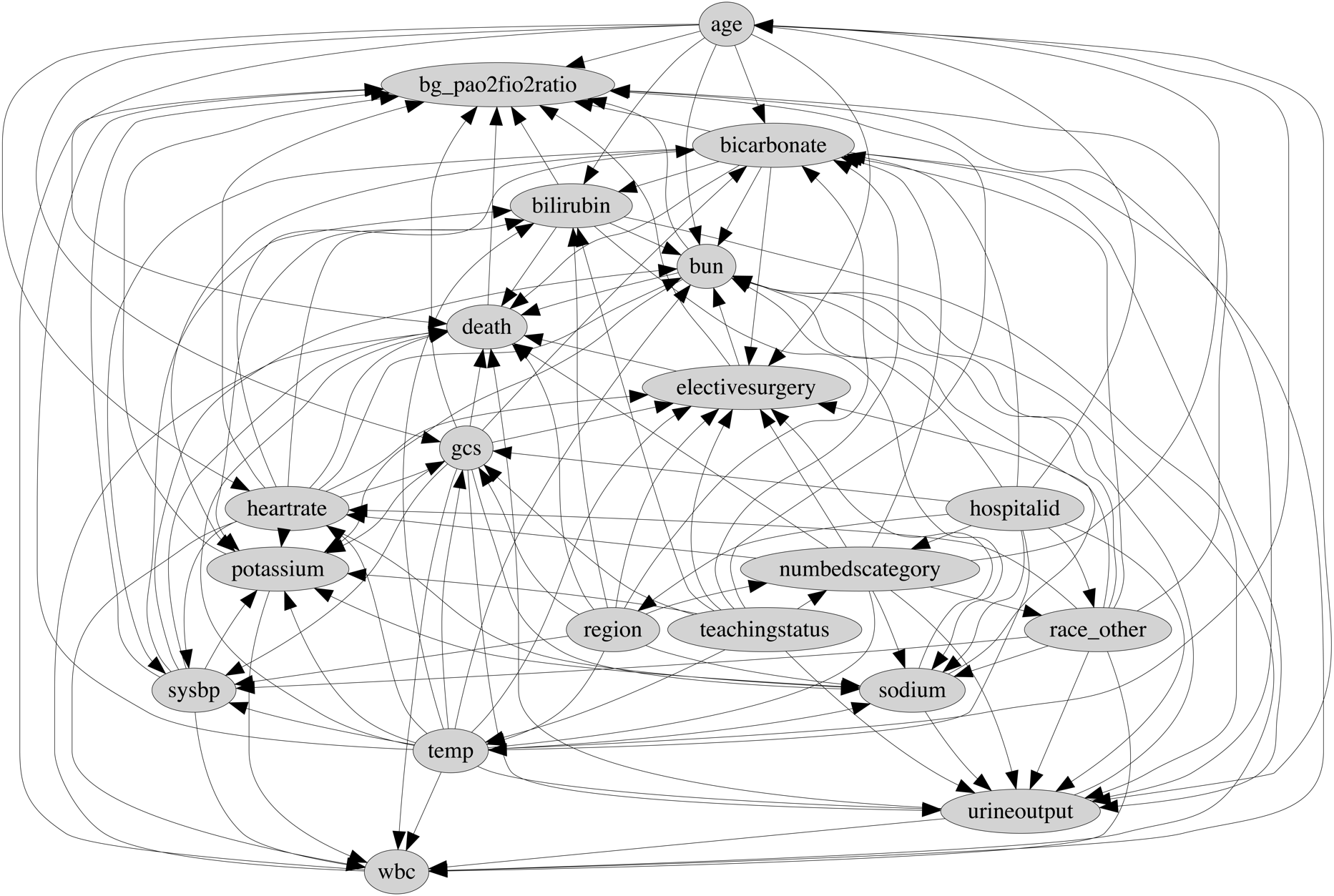

1. Details of pycausal library can be found at <https://github.com/bd2kccd/py-causal>.
2. Tetrad provides a JAVA API for causal discovery. More details can be found at <http://www.phil.cmu.edu/tetrad/>.
